# Wastewater-informed neural compartmental model for long-horizon case number projections

**DOI:** 10.64898/2026.02.10.26345731

**Authors:** Nina Schmid, Nicole Zacharias, Christoph Höser, Johannes Bracher, Jonas Arruda, Cihan Papan, Nico T. Mutters, Jan Hasenauer

## Abstract

Wastewater-based epidemiology provides a low-cost, scalable view of community infection dynamics, but converting these signals into actionable epidemiological insights remains difficult. Mechanistic models offer interpretability, yet, assumptions such as a constant transmission rate limit realism over long simulation horizons and heterogeneous settings. We present a susceptible-exposed-infectious-recovered (SEIR) universal differential equation (UDE) that links wastewater viral loads to case counts and embeds neural networks to represent time-varying parameters. Parameter and prediction uncertainties are quantified using an ensemble method. We assessed the method using newly collected data for Bonn, Germany, as well as published data for five cities in Rhineland-Palatinate, Germany. The proposed approach produces realistic out-of-sample projections of case counts over an up to 50-week test horizon, and it learns city-specific mappings to prevalence that generalise within each location. Compared to SEIR models with fixed transmission rates, the UDE captures non-stationary drivers (policy, behaviour, seasonality) without sacrificing epidemiological structure, while propagating observation and model uncertainty into the projections. Accordingly, the approach facilitates a scalable interpretation and exploitation of wastewater data for the monitoring of infectious diseases.

## Introduction

Public-health decision-making depends on reliable assessments of the burden of infection in the community. Yet, available data streams rarely provide a direct or unbiased view of prevalence^1,2^. The most direct way to estimate infection prevalence is repeated testing of a representative cohort or panel. Such cohort studies (and related sentinel programs) are considered the gold standard because they mitigate biases from care-seeking behaviour and access to diagnostics^3,4^. In practice, however, they are expensive to establish and difficult to sustain at scale, which limits their temporal and geographic coverage. Consequently, many public-health systems rely primarily on routinely reported case counts and related clinical endpoints. These data are readily accessible, but their relationship to true incidence is mediated by testing intensity, eligibility criteria, reporting delays and behavioural effects, producing time-varying under-ascertainment and structural bias^5,6,7^. Wastewater-based epidemiology (WBE) provides a low-cost, scalable view of community infection dynamics, thus offering a compromise^8^. However, converting these signals into actionable epidemiological insights remains difficult. The reason is that the mapping from infections to measured loads is confounded by sewer processes^9^, shedding heterogeneity^10^, post-recovery shedding^11^ and measurement noise. These factors are disease- and site-specific and generally poorly understood.

Modern computational data analysis and mathematical modelling can provide actionable information based on wastewater monitoring. One can distinguish three classes of approaches: (1) deconvolution approaches which reconstruct incidence from wastewater using assumed or inferred shedding load distributions^12,13,14^; (2) mechanistic (often compartmental) transmission models exploiting explicit descriptions of the wastewater observation process to infer epidemiological quantities like prevalence^15,16,17,18,9,19^; and (3) machine learning approaches such as Bayesian statistical learning or gradient boosting trees which map wastewater measurements to clinical case trajectories or growth-rate proxies ^20,9,21^.

Across these approaches, several limitations persist: Deconvolution approaches or approaches relying on transmission modelling can be sensitive to assumptions about shedding timing, prolonged shedding and sewer decay^12,15,17^ and often require rich auxiliary inputs (e.g. detailed mobility or sewer-network information) that are not routinely available^22,23^. Some approaches model time-varying transmission via manually defined breakpoints or other hand-tuned change structures^18^, which limits its scalability. Machine learning models can capture non-linearities, yet may generalise poorly outside their training regime when disease characteristics or behaviour shift^9^. Moreover, uncertainty quantification is often incomplete: measurement noise is frequently propagated, but epistemic uncertainty is less systematically addressed. Finally, many mappings are evaluated over limited time horizons and are using within-period splits rather than truly forward, long-horizon prediction, despite the fact that WBE is most valuable when clinical surveillance becomes less informative over extended time periods.

In this work, we consider the problem of assessing time-dependent disease prevalences and transmission rates based on wastewater data, assuming that case-based reference data (e.g. reported cases and, where available, cohort prevalence) is available only intermittently (Figure 1a). Specifically, during an initial *learning phase*, paired observations from wastewater and a case-based data modality are used to learn a data-driven mapping from observed waste-water signal to the case-based data modality. In a subsequent and longer *inference phase*, the model is driven by wastewater measurements alone to infer latent epidemic quantities. For this setup, we propose an integrative universal differential equation (UDE) framework that couples a transparent mechanistic backbone with learnable components to absorb time-varying and poorly understood processes. UDEs have proven to be well suited for learning latent epidemic drivers from noisy surveillance streams while retaining mechanistic inductive bias^24,25,26,27^. In our framework, the dynamical system encodes infection progression while allowing the transmission and reporting processes to vary smoothly over time via knowledge-constrained neural network parametrizations (Figure 1b). By allowing for smooth changes of the transmission rate, inferring case-based data modalities from wastewater measurements becomes feasible over long horizons. To support public-health situational awareness and response planning, we combine the UDE with ensemble-based uncertainty quantification to provide predictive intervals that reflect both aleatoric and epistemic uncertainty. More generally, this framework offers a flexible foundation for integrative infectious-disease modelling, allowing additional data modalities (e.g. mobility, clinical testing, meteorological covariates) to inform the learned components and enhance interpretability and calibration in the future. We validate the proposed framework on COVID-19 data for up to 50 weeks for the city of Bonn, Germany. Additionally, we showcase its generalisability to other locations and data modalities by applying the model to data of five cities in Rhineland-Palatinate.

**Figure 1:**
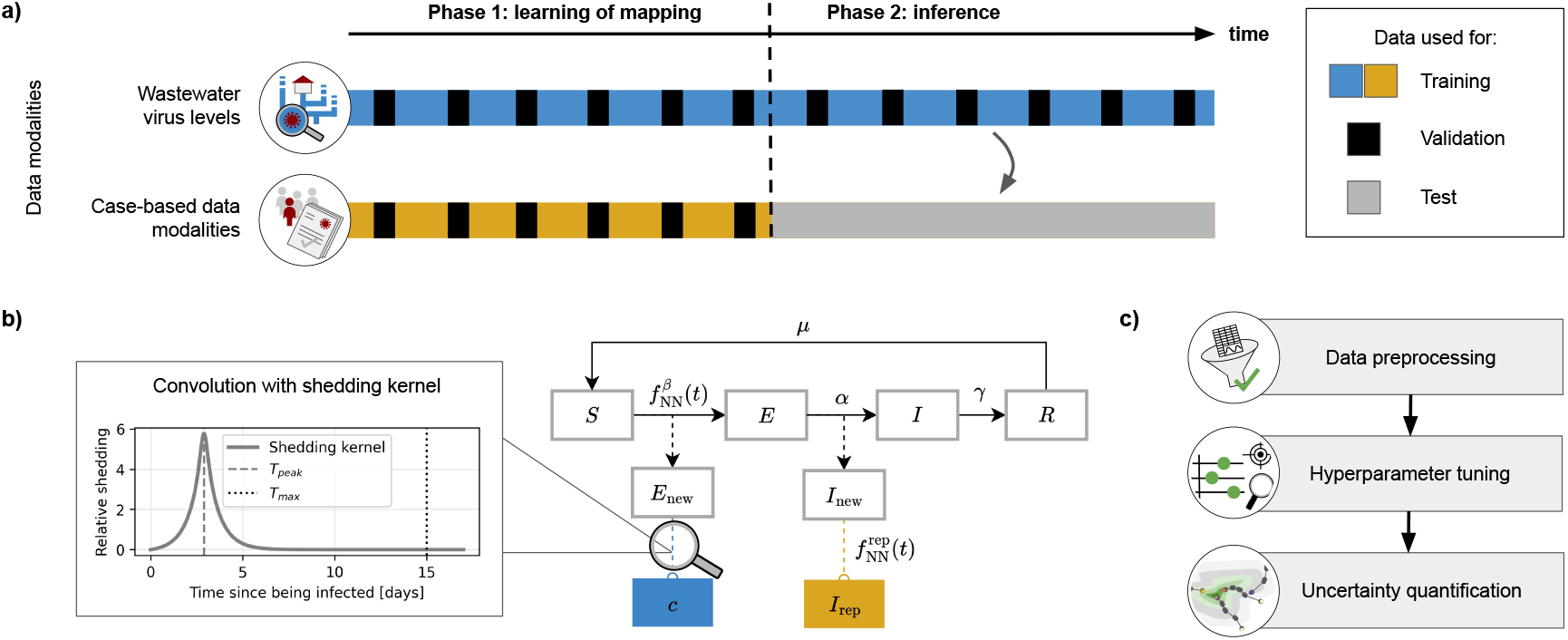
Proposed framework to learn mappings from wastewater data to case-based data modalities. **a)** Indication of data availability during learning and inference phase with highlighting of splitting of respective datasets into training, validation and test. **b)** The transmission dynamics are represented by an SEIR approach, where the transmission rate and the reporting rate are parametrised by neural networks 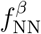 and 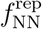. The observation model of the concentration of viral load in wastewater *c* is based on a convolution with a shedding curve. **c)** The complete analysis pipeline starts with data preprocessing, continues with hyperparameter tuning and concludes with ensemble-based uncertainty quantification.

## Results

### A pipeline for learning mappings from wastewater data to case-based data modalities

To learn the mapping from virus levels observed in wastewater to corresponding case-based data modalities such as officially reported cases or prevalence estimates based on cohort testing, we implemented a UDE framework. To calibrate and evaluate the model, available data are split into two subsets, in the following called phases (Figure 1a). In the learning phase, data from both data modalities are necessary to learn the hidden dynamics and mapping. In the inference phase, only wastewater measurements are used to inform the model, while predictions are issued for the case-based data modality.

In order to combine mechanistic knowledge with the flexibility of black-box models, we used a UDE. Specifically, we used a susceptible-exposed-infectious-recovered (SEIR) model as a mechanistic backbone and used literature values for the rates describing disease progression (*α*), recovery (*γ*) and waning immunity (*µ*) (Figure 1b). To allow for long-horizon predictions, we assumed the transmission rate to be time-varying and modelled it using a neural network 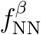 . The shedding of exposed individuals (*E*) into the sewage was modelled using a convolution of newly infected exposed individuals (*E*_new_) with a shedding kernel. The shedding kernel’s parameters are jointly estimated with the remaining parameters under mechanistic constraints, yielding the concentration of viral load in wastewater, *c*. The shape of the shedding kernel is closely related to the shedding curve of infected individuals, yet, due to the simple model structure, it may also capture in-sewer fate and transport processes such as decay and solid-liquid partitioning with settling/resuspension. As not all infected individuals are reported, we introduced a second neural network to learn the time-varying reporting rate 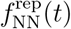, yielding the number of reported cases *I*_rep_. To obtain predictions for the case counts beyond the end time interval available in the learning phase, we adopted the pragmatic assumption that the reporting rate during the inference phase remains constant and is equal to the most recently estimated reporting rate from the learning phase.

We assumed a log-normal noise model for concentration measurements (ensuring a positive and right-skewed distribution) and a negative binomial noise model for officially reported cases (accounting for overdispersion). When fitting a model to data, the noise parameters are optimised jointly with all other parameters, i.e. the parameters of the neural network and the parameters of the shedding curve.

The proposed full modelling pipeline consists of data preprocessing, hyperparameter tuning and uncertainty quantification (Figure 1c). Data preprocessing includes the normalisation of viral loads in wastewater. Although the relative importance of individual hyperparameters varies, hyperparameter tuning has been shown to improve predictive performance and parameter estimation in UDEs^28^. Within our framework, hyperparameters include the number of days it takes on average for a newly infectious person to be reported, the neural network architecture settings and solver settings. Once an appropriate set of hyperparameters is found using a tree-structured Parzen estimator sampler^29^, the framework allows for uncertainty quantification by using multistart ensembles^30^. Specifically, a set of candidate models is defined by sampling many initial parameter values and training one UDE per setting. Due to models getting stuck in local minima and diverging dynamics, fits that do not describe data available during the learning phase sufficiently well have to be filtered out. The resulting ensemble’s variability approximates posterior and predictive uncertainty.

To facilitate the assessment of the proposed framework, we quantified longitudinal measurements of SARS-CoV-2 gene fragments in the sewage in Bonn, together with corresponding variables suitable for normalisation. In addition, we leveraged new case-notification data specific to the exact catchment area of the treatment plant (Figure 2a). To evaluate the framework for five additional cities in Rhineland-Palatinate (Figure 2b) and cohort testing as a case-based data modality, we used a second external dataset that was previously used to infer disease burden from wastewater measurements^19^.

**Figure 2:**
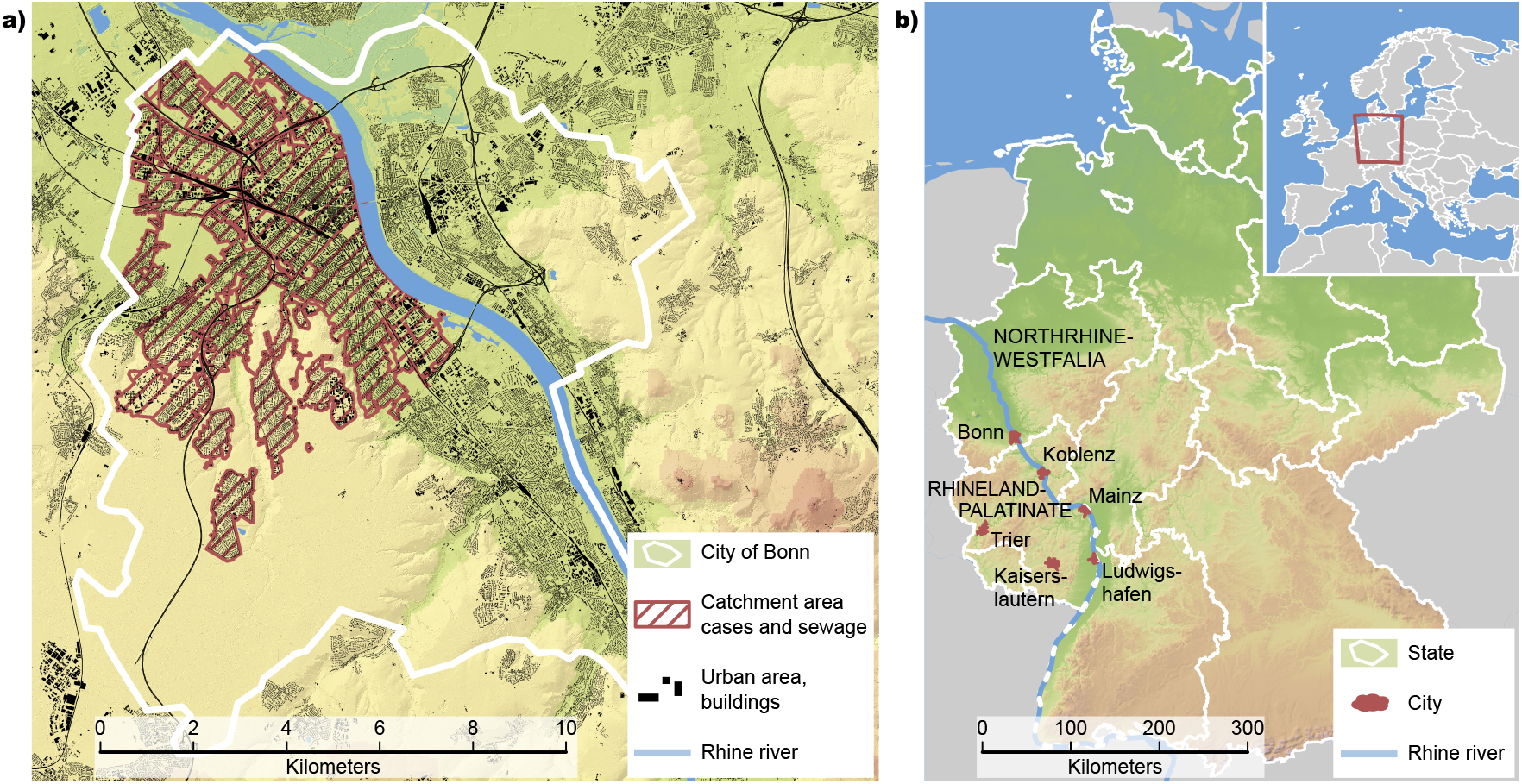
Measurement sites. **a)** Map of the City of Bonn showing the overlapping catchment area of reported cases and sewage data. **b)** Regional context map locating Bonn in North Rhine-Westphalia and Koblenz, Trier, Mainz, Kaiserslautern and Ludwigshafen in Rhineland-Palatinate, with federal state boundaries indicated.

### Wastewater data reliably maps to case counts

To assess the proposed UDE framework, in particular the reliability of the learned mapping, we applied the framework to COVID-19 data from Bonn, Germany. We collected sewage samples between March 2022 and February 2024, quantified the SARS-CoV-2 content and performed normalisation using influent flow volume. In addition, we had access to the officially reported cases in the corresponding catchment area.

We performed comprehensive *in silico* testing of the UDE approach. This revealed that input transformations are vital to the success of the proposed model (Supp. Figure S1). Specifically, we applied an affine transformation to the time variable using fixed reference points, mapping the network input to a scale close to 0. To accommodate an effective learning of fast changing dynamics, we expanded the input to the neural network with Fourier transformations in the time domain^31^. At the same time, we wanted to generate smooth trajectories when data do not suggest differently. This was achieved by implementing a penalty on the magnitude the time derivative of the neural networks takes on. Further, hard constraints on the neural network can ensure that further mechanistic knowledge is not violated. In our case, 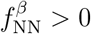 and 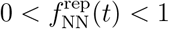 were implemented using the exponential function and the sigmoid function as activation functions of the output layer. For the considered time-period, we assumed that the under-reporting rate is monotonically decreasing. Similar to existing approaches, we enforced this as a hard constraint in the neural network architecture by using monotonically increasing activation functions in combination with a softplus transform of the network weights to ensure their positivity^32^.

With these settings, we applied the optimisation pipeline using a 4, 8, 16, 32 and 50 week evaluation time span. The assessment of the hyperparameter optimisation algorithm revealed that optimisation subsequently improved over the trials, but showed most improvements within the first 100–200 trials (Supp. Figure S2a). Validation and training log-likelihoods yielded similar values across trials (Supp. Figure S2b), indicating a sufficiently flexible model that does not overfit the training data. We found that hyperparameter importances calculated using a functional ANOVA algorithm ^33^ differed between problem scenarios, but learning rates, initial parameter values and the regularisation strength on the transmission rate neural network were frequently among the top five best performing hyperparameters (Supp. Table S1). During ensemble-based multistart optimisation, we observed around 39.4% (minimum 26.5%, maximum 51.1%) of failed optimiser runs (due to memory issues, non-convergence and CPU time limits) across all prediction time frames, indicating that multistart optimisation is vital for achieving acceptable estimates.

The assessment of the predictions across different evaluation time spans revealed that the model could robustly map from viral loads in wastewater to officially reported cases for evaluation time spans of up to 50 weeks (Figure 3a and Supp. Figure S3). The strong performance of our proposed UDE approach stands in contrast to modelling the transmission rate as a time-constant parameter; in that setting, only one of the two data modalities was fitted successfully (Supp. Figure S4). The shorter the horizon in which the UDE-based mapping was used without both data modalities available, the better did predicted and observed case counts match (Figure 3b). Furthermore, the fit diagnostics indicated a systematic trade-off between the two observation modalities. In particular, prediction horizons that showed small negative log-likelihoods associated with the reported cases tended to increase the negative log-likelihoods on the concentration data and vice versa. As concentration observations showed more variation and the variation of the presented model was primarily driven by changes in the transmission rate, this highlights the importance of a suitable neural network regularisation as an important mechanism for balancing fidelity across modalities.

**Figure 3:**
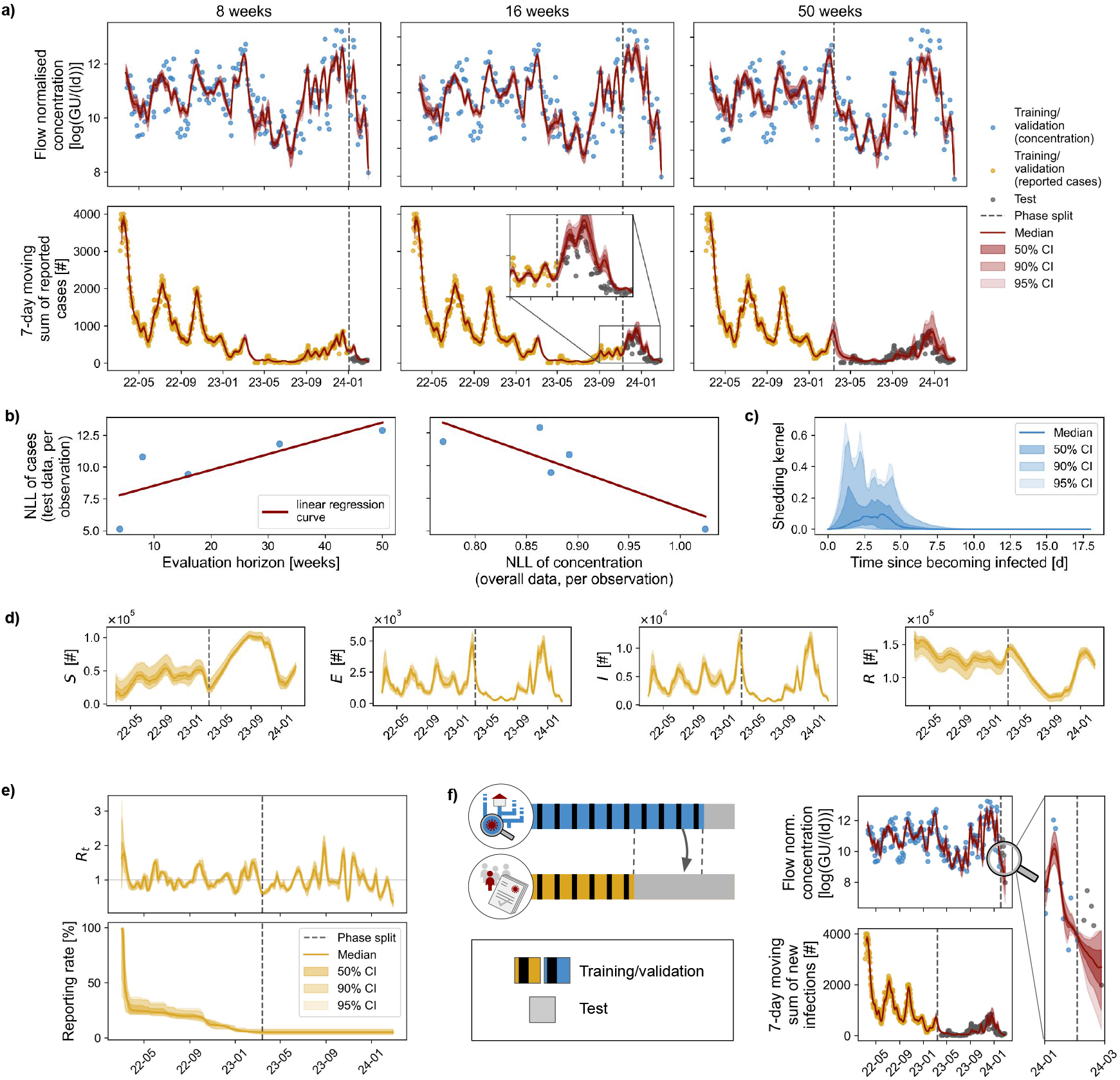
Wastewater data reliably map to case counts. **a)** Fits to concentrations (top) and cases (bottom) across test horizons; confidence intervals (CIs) show ensemble uncertainty of the central estimate (mean/median). **b)** Comparison of negative log-likelihood (NLL) per observation of reported cases on different evaluation horizons, versus evaluation horizon (left) and versus the concentration NLL per observation (overall data) (right) evaluated on the best-validation model from hyperparameter optimisation. **c)** Inferred shedding kernel (this and following subplots are exemplary for the 50-week model). **d)** SEIR hidden-state trajectories. **e)** The effective reproduction number *R*_*t*_ (top) and the reporting rate (bottom) over the simulation time are neural-network-derived quantities. **f)** Data split when concentration data is predicted (left). Fitting results when 3 weeks of the concentration data are assumed unknown (right), with zoom into test region of concentration.

Beyond predictive performance, the model yielded epidemiologically plausible estimates of key model characteristics. The shedding kernel reached its peak value around 2–4 days after susceptibles become infected and its signal to the sewage quickly died out afterwards (Figure 3c). The hidden states underlay fast changing dynamics and had uncertainty bands that were larger than those of the number of reported cases (Figure 3d). With values that fluctuated around 1 (Figure 3e), the estimates of the effective reproduction number laid within a realistic epidemiological range ^34^. In comparison, reported values in the literature often rely on temporally constant assumptions and are inferred from larger, spatially aggregated populations, both of which tend to smooth short-term fluctuations. The reporting rate decays over time until it reaches a median value of 5% (Figure 3e). In the context of COVID-19, this resembles trends reported in literature for 2022–2024^35^.

As the regularisation of 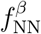 promotes smooth curves of the transmission rate, a prediction into the future for small time horizons is also possible for viral load in wastewater if the dynamics do not change significantly. We showcase this for a prediction horizon of three weeks (Figure 3f). The concentration curve continued the previous weeks’ trend, but epistemic uncertainty rose sharply.

### Cohort testing data allows for assessment of prevalence based on wastewater data

To investigate its generalisability to other cities, we applied our framework to COVID-19 infection data for five other cities in the state of Rhineland-Palatinate, Germany. The data spanned the period between October 2022 and November 2023 and thus contain substantially fewer observations than the data from Bonn. However, in addition to normalised wastewater concentrations and reported cases, cohort testing data collected twice per week were available for these locations from October 2022 and November 2023 (with a 2-month pause starting in June 2023). We used the same feature transforms, hard constraints and regularisation methods as for the Bonn data.

The analysis of data revealed that wastewater viral loads remained broadly stable over time, while reported case counts declined and plateaued near zero. Despite this increased difficulty, the learned models reproduced the reported case dynamics across all cities (Figure 4a), demonstrating accurate wastewater-to-case mapping under non-stationary underreporting.

**Figure 4:**
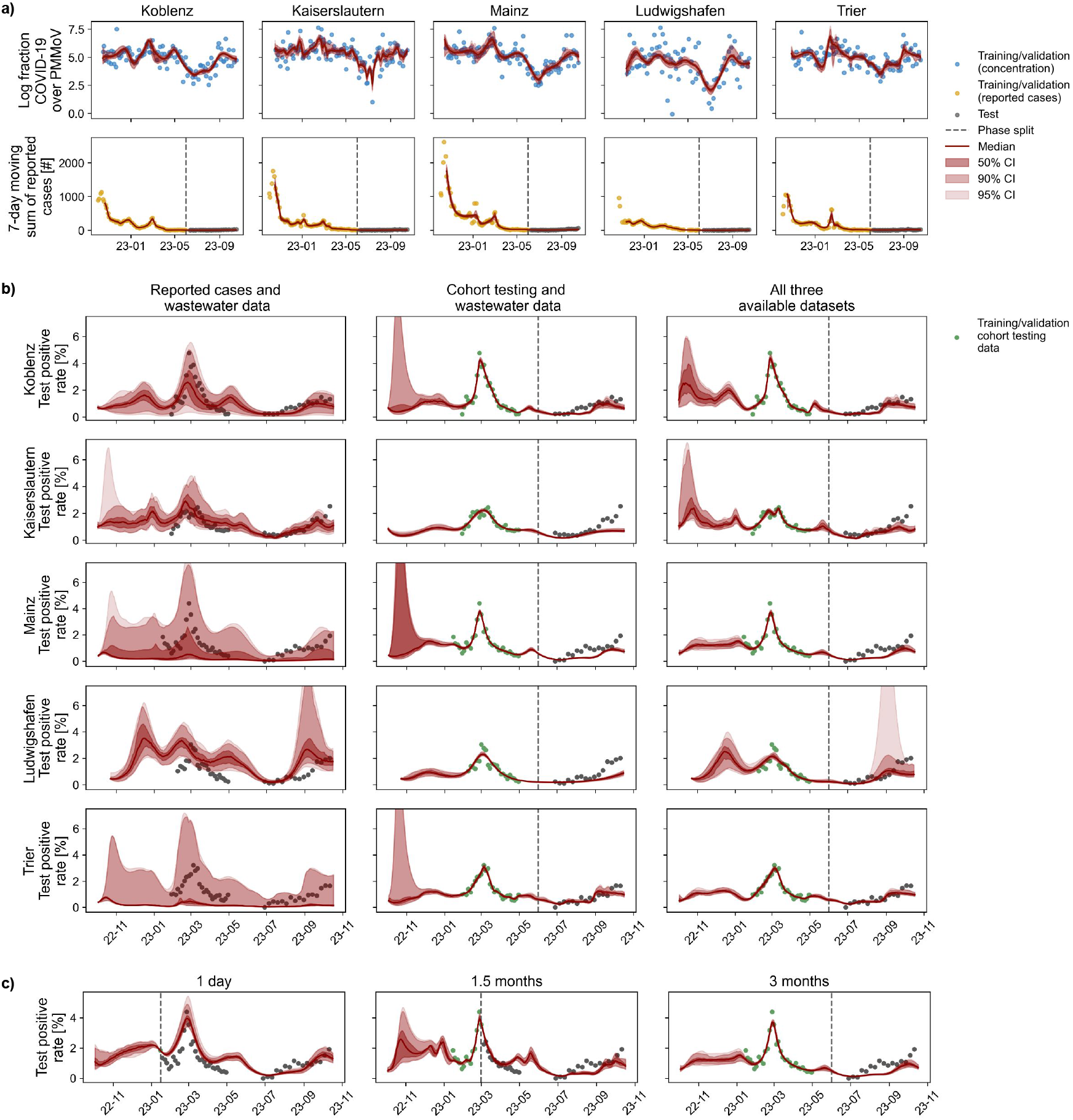
Assessment of prevalence based on wastewater data. **a)** Fits to concentration (top) and case counts (bottom) for five cities in Rhineland-Palatinate using these two data modalities during the learning phase of the mapping. CIs show ensemble uncertainty of the central estimate. **b)** Prevalence estimates under competing objectives using cases and wastewater (left), cohort testing and wastewater (middle) or all three available data sources (right). **c)** Effect of prevalence-data availability on estimation quality (Mainz), holding the case phase split fixed: 1 day (left), 1.5 months (middle) and 3 months (right).

To assess the performance of the framework to estimate prevalence, we utilised the cohort testing data as a proxy of the true underlying prevalence and compared model performance utilising different data sets in the objective function. When using only reported cases and viral load in wastewater as training data, i.e. the setting that we have used for the Bonn dataset, the models had difficulties in simulating the underlying true prevalence values and indicated large uncertainties (Figure 4b). This is plausible, as both data modalities are only qualitatively describing how the underlying number of infections change. Accordingly, we replaced the case-based data modality with cohort test positivity data. For this data, we assumed a binomial noise model, in which we incorporate prior knowledge about sensitivity and specificity of the individual tests. This yielded reasonable fits. Using all three datasets in the objective function further improved the estimation, although for Ludwigshafen we observed larger prediction uncertainties. For all cities, the mean squared errors between median ensemble estimates and observed test positivity rates were smaller and the coverage of the empirical data was increased (Supp. Figure S5).

To understand the influence of the number of cohort testing observations on the quality of the estimation, we reran the analysis using 1 day, 1.5 months and 3 months of cohort testing data during the learning phase while keeping the number of observations on the reported cases unchanged. We observed a good agreement with the data for all three calibration interval scenarios (Figure 4c). The mean squared errors between median ensemble estimates and observed test positivity rates on observations not used during model training were 0.42 (1 day), 0.22 (1.5 months) and 0.28 (3 months). Interestingly, the length of the calibration interval had only a small influence on the prediction accuracy at late time points.

## Discussion

To link wastewater viral loads to case counts for long prediction horizons, we presented a universal differential equation (UDE) framework that preserves a transparent SEIR structure and propagates uncertainty via ensembles. Neural networks were embedded to capture time-varying transmission and reporting processes, so that these quantities function as effective, data-driven summaries of complex and potentially unobserved processes. For instance, the learned transmission rate implicitly reflects and summarises the impact of vaccination uptake, new variants and major social events such as the Carnival or the Christmas periods. By learning an effective shedding kernel, the model aligns wastewater measurements with case-based data modalities.

For our data for Bonn, Germany, the model reconstructed and projected case counts from wastewater over horizons of up to 50 weeks. Across five cities in Rhineland-Palatinate, our model learned city-specific mappings to reported cases; and with additional cohort data, it recovered plausible prevalence trajectories. This indicates that the approach is transferable across locations, provided suitable calibration data are available.

A key observation is that multi-modal data integration is crucial for the identification of process parameters and the prediction of the transmission process. With only wastewater and reported cases, the model captured qualitative trends in latent states, but the absolute level of prevalence remained weakly identified because shedding intensity, reporting and infection burden can trade off against one another. Introducing prevalence measurements from cohort testing clarified this ambiguity: even sparse prevalence data already constrained the model and substantially improved inference of infection levels. This supports the view that WBE should be interpreted as a constituent of an integrative surveillance system, in which high-quality auxiliary data (e.g. cohort testing, serology or hospitalisation rates) help calibrate mechanistic-machine learning hybrids.

The learned components have clear epidemiological interpretations. The inferred shedding kernel peaked shortly after infection, consistent with early high shedding. The effective reproduction number indicated alternating phases of transmission expansion and contraction, while the reporting rate declined to low single-digit percentages, in line with increasing under-ascertainment over 2022–2024. Unlike purely data-driven mappings from wastewater to prevalence^9^, these patterns arose within a constrained SEIR backbone, reducing the risk of overfitting and supporting out-of-sample use. Compared to detailed, site-specific mechanistic models of sewer dynamics^22^ or models with manually specified breakpoints in transmission^18^, our UDE approach automates the learning of non-stationary rates while retaining epidemiological structure and providing a direct, city-level link from wastewater to case-based indicators.

Our framework provides long-horizon case number projections, yet, we assume that the reporting rate remains constant in the prediction phase. This matches the close-to-zero reporting in our setting, but may be implausible where testing campaigns, policy changes or new variants alter case ascertainment more abruptly. This problem could be addressed by learning a functional model for under-reporting based on known factors such as access to testing^5^, stringency of non-pharmaceutical intervention^6^ and public awareness^7^. Our regularisation of the transmission network encodes a smoothness prior: it helps avoid overfitting and improves long-horizon performance, but its optimal strength is problem-specific and our hyperparameter tuning algorithms may only yield a plausible, but not the optimal regularisation strength. Future work should explore alternative regularisation methods that can numerically link this smoothness prior to literature values. Another aspect to consider when applying our framework to new settings is that the model’s shedding kernel conflates individual-level shedding and in-sewer transport and decay into a single effective function, limiting the ability to disentangle host biology from sewer hydraulics. Our approach would also allow one to describe the input-output behaviour of the sewage network using a UDE by introducing additional state variables. This was not done, as we expect the dynamics to be sufficiently fast^22,23^. Furthermore, we assume an endemic setting where key parameters of the disease like the waning immunity rate are already known. By using distributions instead of point estimates of these disease parameters in the ensemble-based uncertainty quantification approach, our framework could be extended to scenarios with uncertainties about all disease parameters. Naturally, this would increase prediction uncertainty and, hence, limit reliable policy guidance. Finally, our approach assumes a period in which wastewater measurements overlap with case or prevalence data; in settings with very short or absent overlap, stronger priors and additional data sources will be needed.

Crucially, the proposed framework provides a practical bridge between mechanistic transparency and data-driven flexibility. The model retains an explicit SEIR backbone while learning time-varying transmission, reporting and shedding relationships directly from the data, thereby aligning wastewater and case modalities. This enables scalable and robust long-horizon projections, whose interpretability is further enhanced with ensemble-based uncertainty quantification. Looking ahead, the neural components of the UDE need not depend on time alone. Future work could let transmission and reporting networks depend on covariates such as mobility, testing volumes, vaccination coverage, meteorological variables or variant frequencies, thereby attributing parts of their temporal variation to observable drivers and improving interpretability. On the application side, the same architecture can be adapted to other pathogens that are trackable in wastewater, with appropriate changes to the compartmental model and priors on time scales (e.g. for influenza, respiratory syncytial virus or hepatitis A). In this way, UDE-based models could form a common backbone for integrative wastewater-based surveillance across pathogens, combining mechanistic insight with the flexibility of scientific machine learning.

## Methods

### Data

In this study, we considered two data collections: One for the city of Bonn (containing wastewater measurements and reported cases) and one for five cities in Rhineland-Palatinate (containing wastewater measurements, reported cases and cohort testing data per city).

For the city of Bonn, 24-hour composite influent wastewater samples were collected twice-weekly at the Bonn wastewater treatment plant and analysed for SARS-CoV-2 N1 and N2 gene fragments as part of the ESI-CorA project^36^. While conceptualising and implementing the proposed framework, the Robert Koch Institute (RKI) harmonised this data alongside wastewater samples of other cities in Germany and made aggregates accessible^36^. The data used in this study is the original data that contains detailed information about different normalisation variables and gene targets. Dilution effects and variability in sewage contributions require normalisation of these measurements to facilitate more robust comparisons across sampling dates and between wastewater treatment plants. The most appropriate normalisation method can be considered site- and sewershed-specific ^37,38,39^. Pepper mild mottle virus (PMMoV) was quantified in all samples as a potential normalisation marker, and information on influent flow volume was collected as an alternative normalisation method. For the Bonn catchment, normalisation by influent flow volume yielded consistent behaviour and was therefore used in all analyses; results are reported as gene units (GU) per liter and day 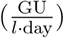. In Bonn, as well as in other cities participating in the surveillance scheme, wastewater SARS-CoV-2 concentrations closely tracked population-level infection trends during both the pandemic and subsequent endemic phases^40^. To assess the proposed framework, we employed a new dataset from the Bonn Health Department (Gesundheitsamt Bonn) containing officially reported case numbers for the exact catchment area of the treatment plant (Figure 2a). The Bonn wastewater and case datasets are provided as Supp. Material.

For Rhineland-Palatinate, we used the wastewater and cohort testing datasets made available as Supp. Information in Mohring et al. (2024) ^19^. Equivalent to the sampling concept in Bonn, 24-hour composite influent samples were collected twice-weekly in each city and analysed for N1 and N2 gene fragments. Both PMMoV as well as flow volume were additionally quantified to allow the comparison of different normalisation methods. In this setting, PMMoV-based normalisation produced more consistent signals across sites^19^ and we therefore used PMMoV-normalised SARS-CoV-2 concentrations. The cohort testing data were collected in the context of the SentiSurv study, in which up to 14,000 participants self-tested for SARS-CoV-2 infection twice per week and reported their COVID-19 status^19,41^. Early in the study, small participant numbers led to large uncertainty in prevalence estimates; for each location, we restricted the analysis to time points with at least 350 participants. Officially reported case numbers for Rhineland-Palatinate cities were obtained from the RKI ^42^. In general, city borders and catchment areas of wastewater treatment plants do not overlap precisely. Hence, our mapping for Rhineland-Palatinate was based on the assumption of relative homogeneity of epidemic dynamics inside the cities and catchment areas.

Supp. Table S2 provides an overview of key characteristics of the different sampling locations and corresponding data. Figure 2b visualises the location of the data sources.

### Epidemic model

We model infection dynamics with a deterministic compartmental system that tracks susceptible, exposed, infectious and recovered individuals in a closed population of size *N* . Let *S*(*t*) (Susceptibles), *E*(*t*) (Exposed), *I*(*t*) (Infectious) and *R*(*t*) (Recovered) denote the corresponding compartment sizes at time *t*. The baseline dynamics follow an SEIR-type ordinary differential equation (ODE) system,

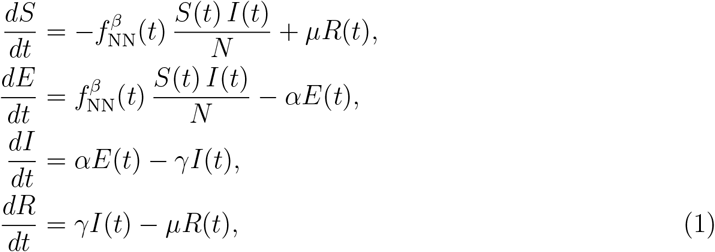

where *α* is the rate at which exposed individuals become infectious, *γ* is the recovery rate and *µ* is the rate at which individuals lose immunity. The time-varying transmission rate ^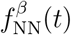^ is represented by a neural network, as described below. *E*(0), *I*(0) and *R*(0) are free parameters that are jointly estimated with all other model components, while *S*(0) = *N* − *E*(0) − *I*(0) − *R*(0). We set 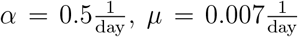 and 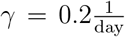, according to typical literature values for the predominant Omicron variant^43,44^.

### Universal differential equation formulation

We embed the mechanistic SEIR structure within a universal differential equation (UDE) framework^27^, in which unknown components are represented by neural networks trained jointly with the ODE parameters. Concretely, we model the effective transmission rate as a feed-forward neural network, whose final activation function is an exponential function to ensure positivity. To provide the network with a flexible yet smooth representation of time, we use *K* Fourier features,

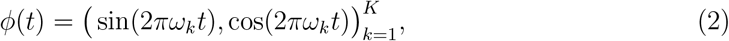

where *ω*_*k*_ are the frequencies and *K* is a hyperparameter. The network takes as input the concatenation of the normalised time *t*_norm_ and *ϕ*(*t*_norm_) and outputs a strictly positive real-valued signal. The normalised time is defined as

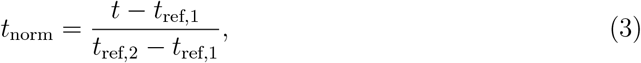

where the reference date *t*_ref,1_ is a time point at the beginning of the time period of interest and *t*_ref,2_ a later time point. In our experiments, *t*_ref,1_ and *t*_ref,2_ were set to the 28th of February 2022 and the 29th of March 2023, respectively.

### Shedding kernel

The shedding kernel *s* is a parametric function that describes how shedding intensity evolves over time since infection, hence, linking the hidden dynamics to measurements of viral load in wastewater. Based on literature research^10^, we assume an exponential rise and decay, whose rise and decay rates *k*_2_ and *k*_3_, as well as the peak time of shedding *T*_peak_ are constrained to plausible values via sigmoid reparametrisation (Supp. Table S3). A sharp logistic weight *w*(*t*) around *T*_peak_ smoothly switches the kernel from the rising branch to the decaying branch, so the overall shape is a smooth, asymmetric peak with data-driven height, timing and skewness. Its amplitude is scaled by *k*_1_, yielding

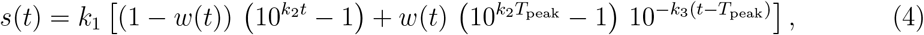

with

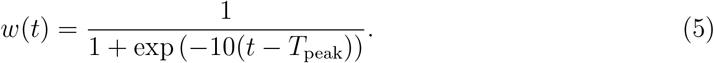

We ensure positivity of *k*_1_ by using an exponential reparametrisation, i.e. we estimate the parameter 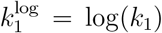. Numerically, all exponentials are clipped in log-space to avoid overflow and ensure stable optimisation.

### Observation models

The model is calibrated using the measured concentration of viral fragments in wastewater *c*_*t*_ and case-based data modalities such as the number of newly reported cases 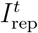 on day *t*. The deterministic model yields the estimates for the number of newly exposed and infectious, *E*_new_(*t*) and *I*_new_(*t*) and estimates for wastewater concentration *c*(*t*) as functions of the latent states:

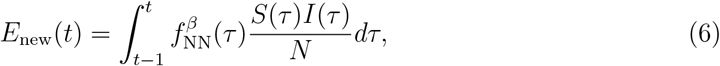

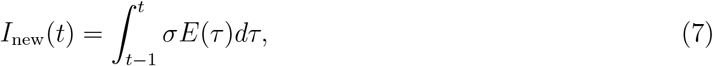

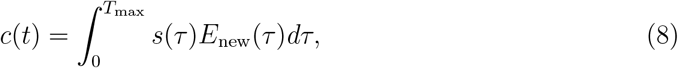

where *T*_max_ is the maximum duration of shedding.

The model accounts for underreporting by estimating the number of reported cases

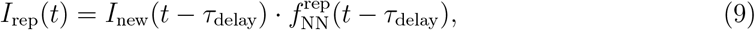

where *τ*_delay_ is an integer reporting delay optimised during hyperparameter optimisation and 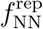 a second feed-forward neural network with a sigmoid activation function in the output layer to generate values between 0 and 1. The input to 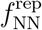 are normalised time points.

For viral load in wastewater, we assume a log-normal noise model with the logarithm of scale as a learnable parameter, thus ensuring positivity and right skew. To account for overdispersion in the discrete counts, we model reported cases by a negative binomial distribution with the variance-mean ratio as a learnable parameter^45^.

For the test data, we use a binomial noise model, where the estimated probability of observing a positive test is given by

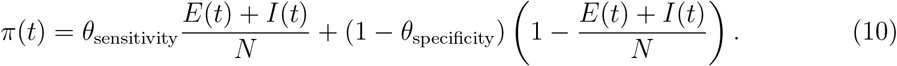

For the test sensitivity *θ*_sensitivity_ and specificity *θ*_specificity_, we use literature values of 0.81 and 1.0, respectively^46^.

All free parameters of the noise models are optimised jointly with the other parameters of the model.

### Optimisation and regularisation

We optimise free parameters *θ* by minimizing the objective function:

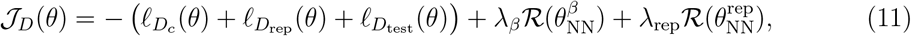

where 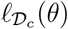 is the log-likelihood of the model predictions with respect to a log-normal noise model and concentration data 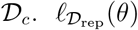 is the log-likelihood with respect to a negative binomial noise model and the data on officially reported cases 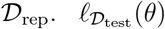 is the log-likelihood with respect to a binomial noise model and the cohort test positivity data 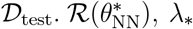 are the regularisation and regularisation strengths, acting on the parameters of the transmission and reporting neural networks.

For a neural network *f*_NN_(*t*; *θ*_NN_) with parameters *θ*_NN_, we penalise the slope of the network, i.e.

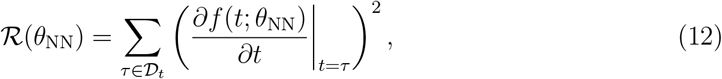

where 𝒟_*t*_ contains all time points of the dataset. Note that different experiments used different case-based data modalities. If the data modality is not available or not used for training, the corresponding training and validation datasets have a cardinality of 0 and the negative log-likelihood is 0.

We infer the model parameters using gradient-based methods, specifically the `Adabelief` optimiser^47^. For this, the dataset is split into training, validation and test sets. The train-validation split is implemented randomly with 1/8 being used as validation data, the test set is defined based on the “phase split date”, i.e. the date after which the model needs to map from wastewater data to case-based data modalities without relying on case observations. Supp. Table S4 lists the phase split dates used in each experiment. We use a learning rate scheduler with four different learning rate values, where its values and the number of optimisation steps per learning rate setting are determined via hyperparameter tuning.

### Hyperparameter tuning

The model and the optimisation procedure rely on several tuneable hyperparameters. Details of the neural network architecture (depth, width, activation function, number of Fourier frequencies *K*), the numerical solver, its tolerances, the number of days of reporting delay and the hyperparameter of the shedding (*T*_max_) specify the model. Hyperparameters such as the learning rates, number of optimiser steps and regularisation strengths define the optimisation process. We perform hyperparameter optimisation using `Optuna` ^48^ and its implementation of a tree-structured Parzen estimator sampler^29^. We used the same hyperparameter search space for all problems (see Supp. Table S5 for details), allowed for the same CPU time and memory and run ≈ 500 trials per problem setup. Given the objective of finding a set of hyperparameters that *can* describe the data, we use the same train-validation split. The best-performing configuration is used for the ensemble-based uncertainty quantification, where we use one train-validation split per potential ensemble member.

### Uncertainty quantification

We estimate epistemic uncertainty of the UDE with an ensemble based method^30^. The variability among the ensemble members reflects randomness in the train-validation split and the initial parameter values. For each problem, we set a maximum computation time of 3200 CPU hours to generate up to 10,000 potential ensemble members; the sampling space is summarised in Supp. Table S6. Due to models getting stuck in local minima and diverging dynamics, not all models yield acceptable fits to the data. The resulting multi-objective setting therefore requires an explicit preselection of ensemble candidates. Specifically, we retain only those models whose negative log-likelihood lies within the top 25% (i.e. below the 25th percentile) for each respective dataset, while forming the final ensemble by selecting the 5% of models achieving the lowest negative log-likelihood on the combined training and validation sets.

### Implementation details

All models are implemented in `JAX`^49^, using `Equinox`^50^ for neural network modules and `Diffrax` for numerical solution of the ODEs^51^. Optimisation is carried out with `Optax` ^52^. The experiments were conducted on Intel Xeon “Sapphire Rapids” 2.10GHz CPUs.

All code used in this study is available on GitHub at https://github.com/schminin/wastewater informed neural compartemental model.

All geospatial base layers used to generate Figure 2 are publicly available from their respective providers: OpenStreetMap (https://www.openstreetmap.org/), BKG Geodatenzentrum (https://gdz.bkg.bund.de/), Geobasisdaten und -dienste der Bezirksregierung Köln, Geobasis NRW (https://www.bezreg-koeln.nrw.de), Natural Earth (https://www.naturalearthdata.com) and CLC Copernicus Land Cover (https://sdi.eea.europa.eu/catalogue/copernicus). The Bonn catchment boundary dataset was obtained from the City of Bonn (2022) and is not publicly available; access may be granted by the City of Bonn and/or is available from the corresponding author upon reasonable request and subject to permission.

## Supporting information

Supp. Data for the City of Bonn

## Data Availability

All data produced in the present work are contained in the Supplementary Material, already published data that were used for this work are available online (details can be found in the manuscript).

https://github.com/schminin/wastewater_informed_neural_compartemental_model

## Acknowledgements

N.S., J.H. and J.A. received funding by the European Union via ERC grant INTEGRATE (grant no 101126146). N.S. and J.H. received further funding from the German Federal Ministry of Research, Technology and Space (BMFTR) (INSIDe - grant number 031L0297A). J.H. further received funding from the Deutsche Forschungsgemeinschaft (DFG, German Research Foundation) under Germany’s Excellence Strategy (EXC 2047 - 390685813, EXC 2151 - 390873048) and by the University of Bonn (via the Schlegel Professorship). J.B. was supported by the German Research Foundation (DFG), project 512483310. N.Z., C.H., C.P. and N.T.M. were funded as part of the EU project ESI-CorA (Detection of SARS-CoV-2 in wastewater), funding was provided through Karlsruhe Institute of Technology, with EU funds (No. 060701/2021/864650/SUB/ENV.C2) passed on to the Institute for Hygiene and Public Health, University Hospital Bonn.

We thank the Bonn Health Department (Gesundheitsamt Bonn) for providing officially reported case counts for the specific catchment area and the civil engineering office of Bonn (Tiefbauamt Bonn) for providing wastewater samples. The authors gratefully acknowledge the access to the Marvin cluster of the University of Bonn.

## Author contributions

N.S.: Conceptualisation, methodology, software, formal analysis, visualisation, investigation, writing - original draft, writing - review & editing; N.Z.: Conceptualisation, data curation, writing - review & editing; C.H.: Conceptualisation, data curation, visualisation, writing - review & editing; J.B.: Conceptualisation, writing - review & editing; J.A.: Conceptualisation, writing - review & editing; C.P.: Conceptualisation, writing - review & editing; N.T.M.: Conceptualisation, writing - review & editing, funding acquisition; J.H.: Conceptualisation, methodology, writing - review & editing, supervision, funding acquisition.

## Competing Interests

The authors declare that they have no competing interests.

## Supporting information

### Supplementary Figures

**Figure S1:**
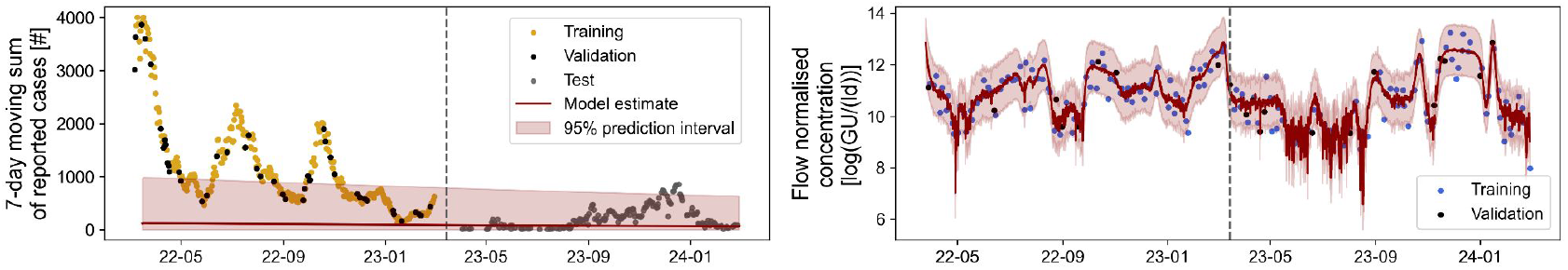
Fit in absence of input normalisation. The best performing model after hyperparameter optimisation is not capable of fitting any data modality sufficiently well. The case data are not fitted at all, while the fit on the concentration data does not yield a sufficiently smooth curve. To generate this plot we used the Bonn dataset, a 50-week prediction horizon and apart from a missing input normalisation the same setup as used for the framework’s application to the Bonn data. The bands indicate the 95% prediction interval of the UDE, hence, visualise the aleatoric uncertainty of the model.

**Figure S2:**
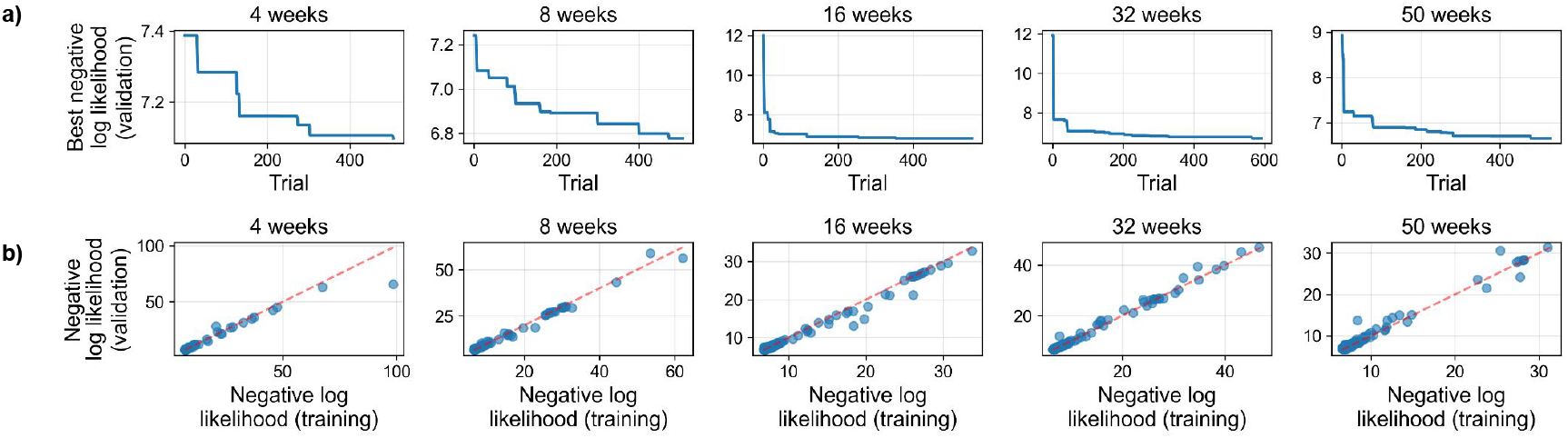
Hyperparameter optimisation. **a)** Convergence of the tree-structured Parzen estimator sampler over the hyperparameter trials. Shown is the best negative log-likelihood value (on validation data) so far at a given trial number. **b)** The negative log-likelihood of validation vs. training data. To investigate only converged fits, only optimiser runs with a validation loss smaller than 10 times the minimal validation loss are displayed.

**Figure S3:**
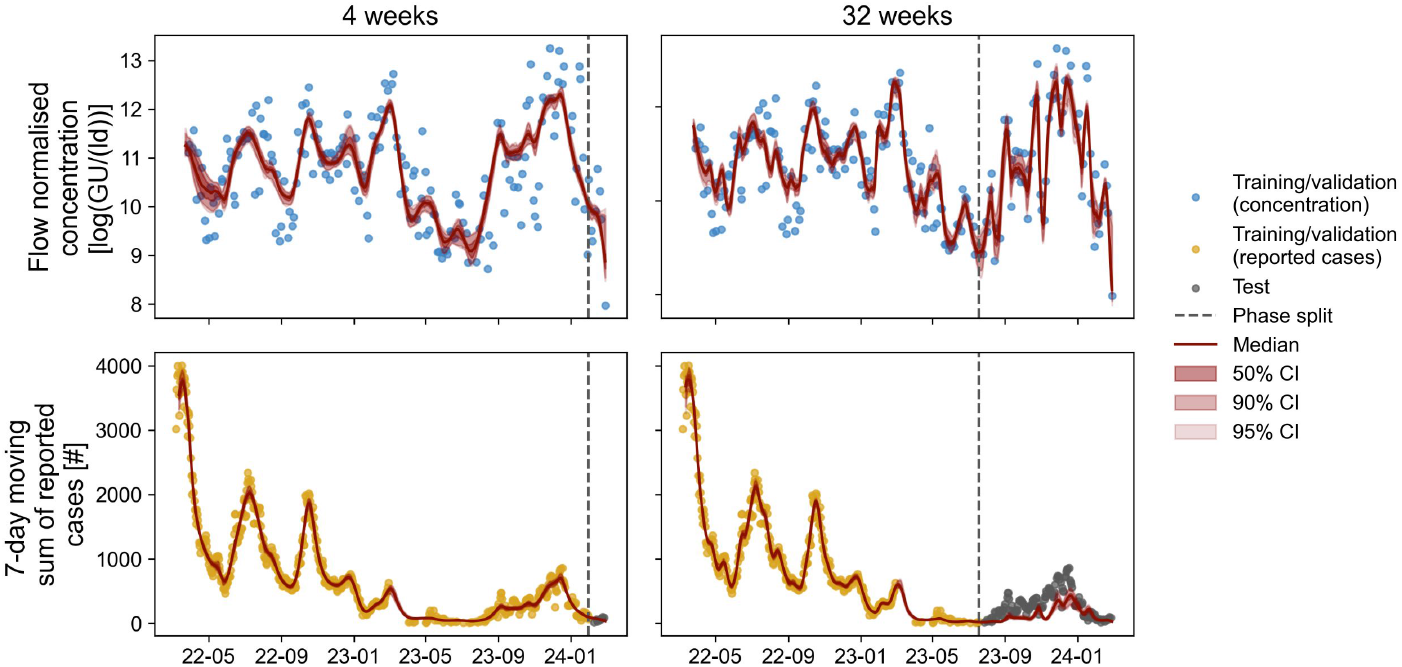
Fitting results. Fitting results for the concentration data (top row) and case counts (bottom row) for different test horizons.

**Figure S4:**
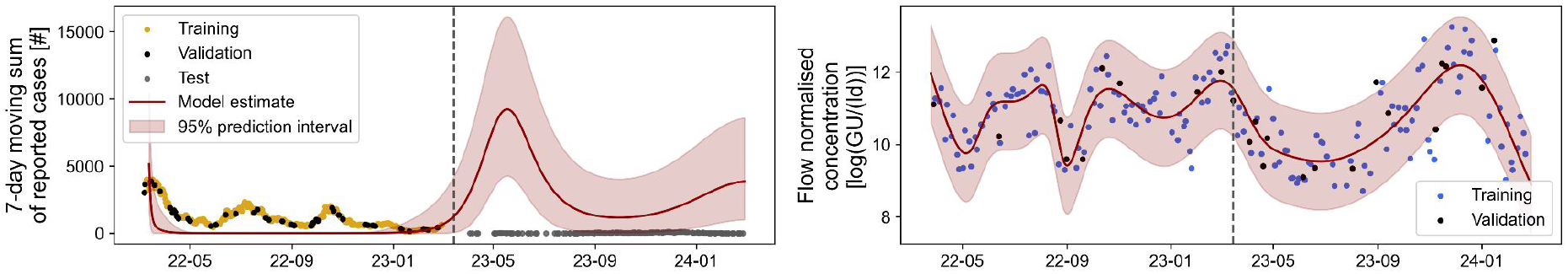
Fit assuming constant transmission rate. The best performing model after hyperparameter training is not capable of fitting both data modalities sufficiently well. The bands indicate the 95% prediction interval of the UDE, hence, visualise the aleatoric uncertainty of the model.

**Figure S5:**
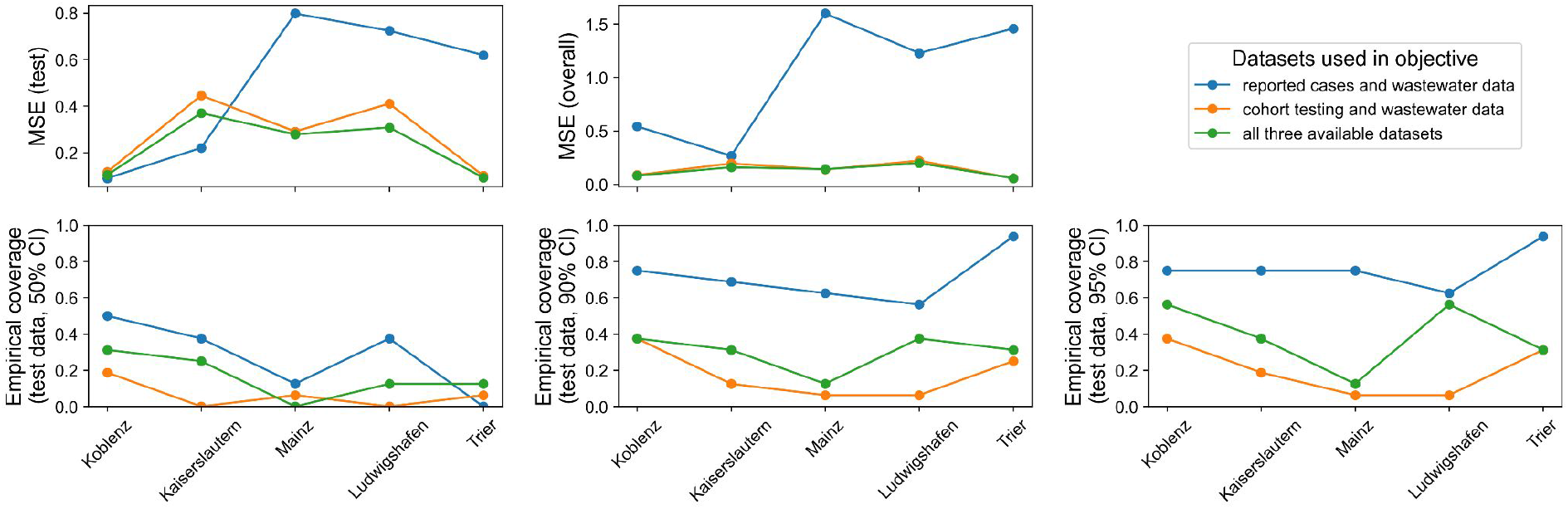
Evaluation of different objectives. The mean squared error (MSE) on the test set (i.e. observed cohort testing after the phase split date) compared to median ensemble estimates and all available cohort testing observations for different cities and datasets used in the objective formulation (top row). The empirical coverage of test data for different confidence intervals of the ensemble based estimate of the test positive rate (bottom row).

### Supplementary Tables

**Table S1:** Hyperparameter importance. Using a functional ANOVA algorithm ^33^, we evaluate hyperparameter importance based on the results of the hyperparameter optimisation performed for an evaluation horizon of 4, 8, 16, 32 and 50 weeks. Displayed are the 5 most important hyperparameters (ordered declining according to their importance). `step*` describes the number of optimiser steps in phase `*` of the learning rate scheduler, `lr_step*` the corresponding learning rate. `*_init` corresponds to initial mechanistic parameter values and `reg_norm` is the regularisation strength of the transmission rate neural network, *λ*_*β*_.

| rank | 4 weeks | 8 weeks | 16 weeks | 32 weeks | 50 weeks |
| --- | --- | --- | --- | --- | --- |
| 1 | step3 | lr_step1 | reg_norm | lr_step1 | lr_step1 |
| 2 | lr_step1 | k3_init | step3 | reg_norm | width_size |
| 3 | width_size | step2 | T_peak_init | k2_init | activation |
| 4 | init_sigma_C | reporting_delay | step4 | lr_step2 | rtol |
| 5 | step2 | reg_norm | width_size | k1_init | step4 |

**Table S2:** Overview of sampling locations.

| City | Catchment areas' population size | Case and wastewater data are based on same catchment | Available case-based data modalities |
| --- | --- | --- | --- |
| Bonn | 176,187 | yes | reported cases |
| Mainz | 220,552 | no | reported cases, cohort testing |
| Trier | 112,195 | no | reported cases, cohort testing |
| Koblenz | 115,268 | no | reported cases, cohort testing |
| Kaiserslautern | 108,216 | no | reported cases, cohort testing |
| Ludwigshafen | 174,265 | no | reported cases, cohort testing |

**Table S3:** Constraints of the shedding kernel’s parameters.

| Parameter name | Lower limit | Upper limit |
| --- | --- | --- |
| $k_2$ | $0.6 \frac{1}{\text{day}}$ | $2.5 \frac{1}{\text{day}}$ |
| $k_3$ | $0.15 \frac{1}{\text{day}}$ | $2.0 \frac{1}{\text{day}}$ |
| $T_{\text{peak}}$ | 1 day | 5 days |

**Table S4:** Phase split dates and objective functions of the different experiments using the proposed framework. For the setting annotated with (*) we conducted a second experiment, where we also introduce a phase split date for concentration (yielding concentration predictions for 3 weeks).

| City | phase split date<br>(reported cases) | phase split date<br>(test positive rate) | datasets used for objective |
| --- | --- | --- | --- |
| Bonn | 2024-01-31 | - | reported cases & wastewater |
| Bonn | 2024-01-03 | - | reported cases & wastewater |
| Bonn | 2023-11-08 | - | reported cases & wastewater |
| Bonn | 2023-07-19 | - | reported cases & wastewater |
| Bonn (*) | 2023-03-15 | - | reported cases & wastewater |
| Mainz | 2023-06-01 | 2023-06-01 | all three |
| Mainz | 2023-06-01 | 2023-03-01 | all three |
| Mainz | 2023-06-01 | 2023-01-16 | all three |
| Trier | 2023-06-01 | 2023-06-01 | all three |
| Ludwigshafen | 2023-06-01 | 2023-06-01 | all three |
| Kaiserslautern | 2023-06-01 | 2023-06-01 | all three |
| Koblenz | 2023-06-01 | 2023-06-01 | all three |
| Mainz | - | 2023-06-01 | wastewater & cohort testing |
| Trier | - | 2023-06-01 | wastewater & cohort testing |
| Ludwigshafen | - | 2023-06-01 | wastewater & cohort testing |
| Kaiserslautern | - | 2023-06-01 | wastewater & cohort testing |
| Koblenz | - | 2023-06-01 | wastewater & cohort testing |
| Mainz | 2023-06-01 | - | reported cases & wastewater |
| Trier | 2023-06-01 | - | reported cases & wastewater |
| Ludwigshafen | 2023-06-01 | - | reported cases & wastewater |
| Kaiserslautern | 2023-06-01 | - | reported cases & wastewater |
| Koblenz | 2023-06-01 | - | reported cases & wastewater |

**Table S5:** Hyperparameter search space.

| Hyperparameter | Type / distribution | Range / values |
| --- | --- | --- |
| Neural network width | integer, uniform | [4, 32] |
| Neural network depth | integer, uniform | [1, 3] |
| Activation function | categorical | {tanh, sigmoid} |
| Number of Fourier features | integer, uniform | [0, 3] |
| Initial $T_{\max}$ | integer, uniform | [12, 28] |
| Initial $T_{\text{peak}}$ | float, uniform | [1, 5] |
| Initial $k_1$ | float, log-uniform | [0.01, 40] |
| Initial $k_2$ | float, uniform | [0.6, 2.5] |
| Initial $k_3$ | float, uniform | [0.15, 2.0] |
| Initial variance-mean ratio (case noise model) | float, log-uniform | [0.01, 10] |
| Initial $\sigma^2$ (concentration noise model) | float, log-uniform | [$10^{-3}$, 1] |
| Reporting delay [days] | integer, uniform | [0, 10] |
| ODE solver | categorical | {Dopri5, Tsit5} |
| Relative tolerance of ODE solver | float, log-uniform | [$10^{-4}$, $10^{-2}$] |
| Absolute tolerance of ODE solver | float, log-uniform | [$10^{-5}$, $10^{-3}$] |
| Learning rate, phase 1 | float, log-uniform | [$10^{-4}$, $10^{-1}$] |
| Learning rate, phase 2 | float, log-uniform | [$10^{-5}$, $10^{-2}$] |
| Learning rate, phase 3 | float, log-uniform | [$10^{-5}$, $10^{-2}$] |
| Learning rate, phase 4 | float, log-uniform | [$10^{-5}$, $10^{-2}$] |
| Optimisation steps, phase 1 | integer, uniform | [500, 3000] |
| Optimisation steps, phase 2 | integer, uniform | [500, 3000] |
| Optimisation steps, phase 3 | integer, uniform | [500, 3000] |
| Optimisation steps, phase 4 | integer, uniform | [500, 4000] |
| Regularisation strength<br>for transmission model | float, log-uniform | [$10^{-5}$, $10^{-3}$] |
| Regularisation strength<br>for under-reporting model | float, log-uniform | [$10^{-5}$, $10^{-3}$] |

**Table S6:** Multi-start sampling settings. The superscript ^“HP”^ references a result from hyperparameter optimisation.

| Parameter | Type / distribution | Range / values /distribution |
| --- | --- | --- |
| Initial $T_{\text{peak}}$ | float, uniform | $[0, 5]$ |
| Initial $k_1$ | float, log-uniform | $[0.01, 40]$ |
| Initial $k_2$ | float, uniform | $[0.6, 2.5]$ |
| Initial $k_3$ | float, uniform | $[0.15, 2.0]$ |
| Initial $E(0)$ | float, normal | $\mathcal{N}(E^{\text{HP}}(0), 0.1E^{\text{HP}}(0))$ |
| Initial $I(0)$ | float, normal | $\mathcal{N}(I^{\text{HP}}(0), 0.1I^{\text{HP}}(0))$ |
| Initial $R(0)$ | float, normal | $\mathcal{N}(R^{\text{HP}}(0), 0.1R^{\text{HP}}(0))$ |
| Reporting delay | integer, uniform<br>with constraint | $\pm 2$ days of $\tau_{\text{delay}}^{\text{HP}}, > 0$ |
| Neural network<br>weight initialization | float, (truncated)<br>normal | $\mathcal{N}(1, \frac{1}{\sqrt{n_{\text{av}}}})$, with $n_{\text{av}}$ being the arithmetic<br>average of the numbers of inputs and outputs;<br>absolute values of the samples are truncated<br>at 2 standard deviations before scaling |

## Notes

### Competing Interest Statement

The authors have declared no competing interest.

### Author Declarations

This study used both previously published data, as well as new data. For Rhineland-Palatinate, we used the wastewater and cohort testing datasets made available as Supp. Information in Mohring et al. (2024). Officially reported case numbers for Rhineland-Palatinate cities were obtained from the Robert Koch Institute. For Bonn, the Bonn Health Department (Gesundheitsamt Bonn) provided officially reported case counts for the specific catchment area and the civil engineering office of Bonn (Tiefbauamt Bonn) for provided wastewater samples, which we then analysed for SARS-CoV-2 gene fragments alongside normalisation variables. This data has been de-identified prior to use in the study. An Ethics approval was not necessary.

